# Estimating Cases of COVID-19 from Daily Death Data in Italy

**DOI:** 10.1101/2020.03.17.20037697

**Authors:** Ali Raheem

## Abstract

COVID-19 is an emerging infectious disease which has been declared a pandemic by the World Health Organisation. Due to limited testing capacity for this new virus, variable symptomatology the majority of infected showing non-specific mild or no symptoms it is likely current prevalence data is an underestimate.

**Methods:** We present an estimate of the number of cases of COVID-19 compared to the number of confirmed case in Italy based on the daily reported deaths and information about the incubation period, time from symptom onset to death and reported case fatality rate.

**Results:** Our model predicts that on the 31^st^ of January 2020 when the first 3 infected cases had been identified by Italian authorise there were already nearly 30 cases in Italy, and by the 24th of February 2020 only 0.5% cases had been detected and confirmed by Italian authorities. While official statistics had 132 confirmed case we believe a more accurate estimate would be closer to 26000. With a case-doubling period of about 2.5 days.

## Introduction

COVID-19 has now been declared a pandemic by the World Health Organisation. Caused by a betacoronvirus virus SARS-CoV2 which is related to the SARS and MERS viruses. This disease appears to have a case fatality rate of approximated 2–15%. However there have been a wide variety in reported proportion of cases that are asymptomatic or only show mild non-specific symptoms. Making it difficult to estimate the number of cases of COVID-19 without widespread testing which has not yet been implemented in any country. While frequently case fatality rate has been calculated by comparing spot dates in 24 hours to spot confirmed cases in the same 24 hours, this is inaccurate (1). Accurately estimating the prevalence of COVID-19 will allow organisations to make better informed decisions to control COVID-19. The majority of testing is currently being done with Reverse Transcription Polymerase Chain Reaction (RT-PCR) methodology which can be done in real-time at some labs, confirmation may use Nucleic Acid Amplification Test (NAAT) methodology (2). As infections outstrip testing capacity we risk infectious casing slipping through our fingers, without confirmation it’s possible some quarantine subjects will be less willing to self-isolate increasing the likelihood of further spread.

## Methods

We used a linear retrospective model to estimate past point prevalence using daily number of report deaths. This model required us to calculate a nominal time to death from infection. We based this value on data available from the World Health Organisation. The WHO report time from onset of symptoms to death of about 2 weeks (WHO, 2020). Additionally, the mean incubation period has been reported to be 6.4 days (3). We estimate that deaths on a given day should correlate with infections 3 weeks prior and use this with daily reported deaths to estimate the spot prevalence in the past. We obtained data on daily deaths and past cases from the European Centre for Disease Prevention and Control (4) cross referenced for accuracy with data from the World Health Organisation (5).

Data processing was carried out with R, the Juptyer notebook and Tidyverse software suites on a Debian 9.0 Stretch using the latest Jupyter/r-notebook docker image (jupyter/r-notebook:15a66513da30) (6) (7).

Using a case mortality of 7% was used based on the recent estimates.

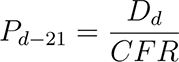

CFR Where *P* is prevalence, *D* is deaths reported, *CFR* is the case fatality rate, and *d* is time in days. Using a case fatality rate of 7% we have:

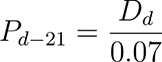

Due to our model varying with the reciprocal of *CFR*, we expect little impact on *CFR* error on cases in the range of *CFR* reported, see Figure 1 for a graphical depiction.

The following data was generated using the reported number of deaths per day and new confirmed cases. From this a cumulative cases and cumulative deaths data was calculated and used to calculate the point prevalence according to the formula described above.

Using data up to the 16^th^ of March 2020 we can estimate the point prevalence 21 days into the past (the 2^th^ 4 of February 2020). This value is subject to the inevitable jitter in deaths per day due to COVID-19, it therefore should be used to guide a trend line before interpretation.

**Figure 1:**
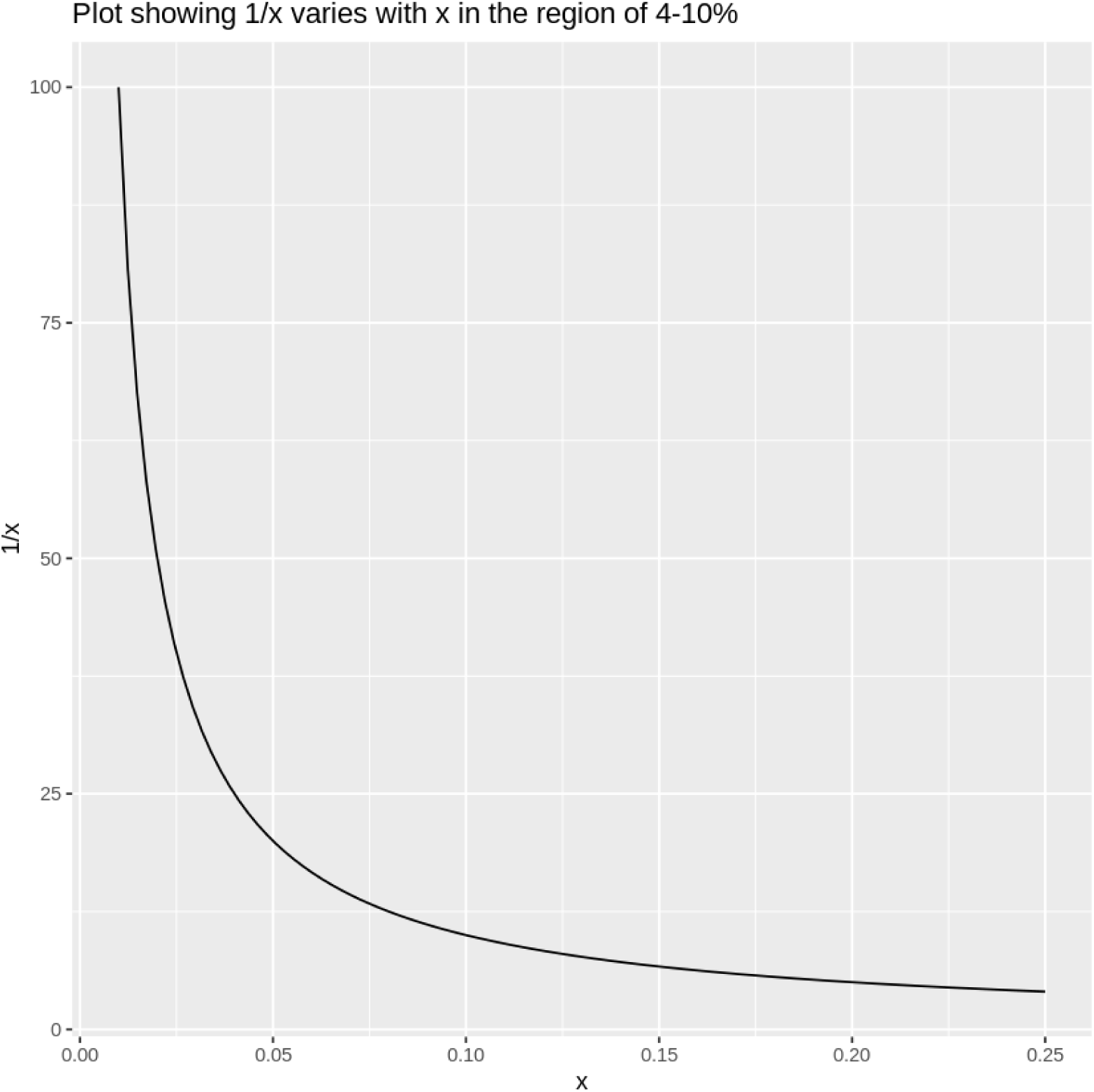
Plotting 1/x, notice the relative flatness from 0.04 up showing that the model will be robust to varibality in *CFR*.

## Results

Figure 2 summaries the results graphically, the full results can be reviewed in the supplementary materials section in Table 1.

**Figure 2:**
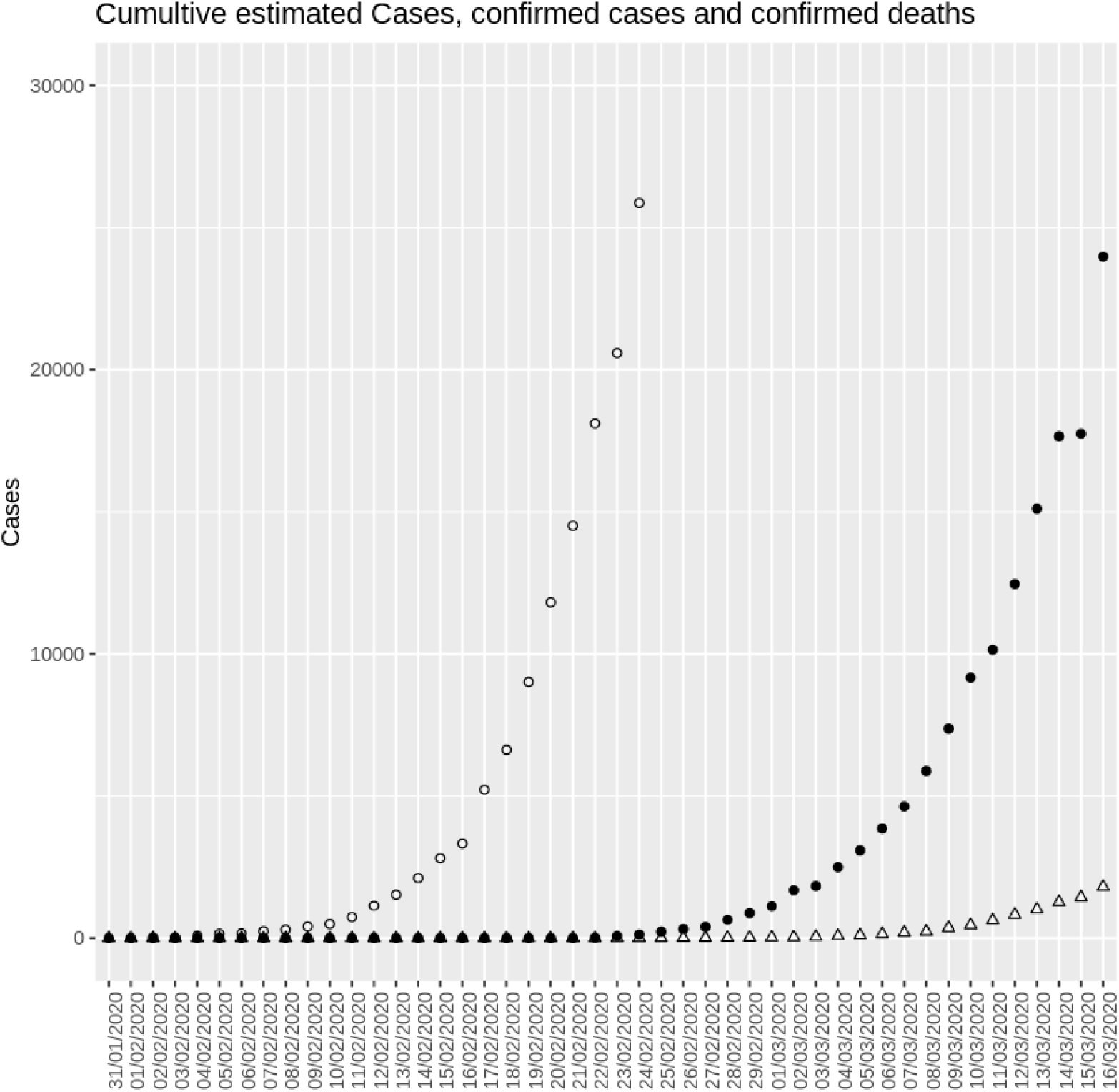
Plotting Estimated Prevalence (circles), Confirmed Cases (filled triangles), Reported Deaths (triangles)

**Table 1:** Raw results data

| Date | New Cases | New Deaths | Total Cases | Total Deaths | Est Total Cases |
| --- | --- | --- | --- | --- | --- |
| 31/01/2020 | 3 | 0 | 3 | 0 | - |
| 01/02/2020 | 0 | 0 | 3 | 0 | - |
| 02/02/2020 | 0 | 0 | 3 | 0 | 28.57143 |
| 03/02/2020 | 0 | 0 | 3 | 0 | 28.57143 |
| 04/02/2020 | 0 | 0 | 3 | 0 | 85.71429 |
| 05/02/2020 | 0 | 0 | 3 | 0 | 157.14286 |
| 06/02/2020 | 0 | 0 | 3 | 0 | 171.42857 |
| 07/02/2020 | 0 | 0 | 3 | 0 | 242.85714 |
| 08/02/2020 | 0 | 0 | 3 | 0 | 300.00000 |
| 09/02/2020 | 0 | 0 | 3 | 0 | 414.28571 |
| 10/02/2020 | 0 | 0 | 3 | 0 | 500.00000 |
| 11/02/2020 | 0 | 0 | 3 | 0 | 742.85714 |
| 12/02/2020 | 0 | 0 | 3 | 0 | 1142.85714 |
| 13/02/2020 | 0 | 0 | 3 | 0 | 1528.57143 |
| 14/02/2020 | 0 | 0 | 3 | 0 | 2114.28571 |
| 15/02/2020 | 0 | 0 | 3 | 0 | 2814.28571 |
| 16/02/2020 | 0 | 0 | 3 | 0 | 3328.57143 |
| 17/02/2020 | 0 | 0 | 3 | 0 | 5228.57143 |
| 18/02/2020 | 0 | 0 | 3 | 0 | 6628.57143 |
| 19/02/2020 | 0 | 0 | 3 | 0 | 9014.28571 |
| 20/02/2020 | 0 | 0 | 3 | 0 | 11814.28571 |
| 21/02/2020 | 0 | 0 | 3 | 0 | 14514.28571 |
| 22/02/2020 | 14 | 0 | 17 | 0 | 18114.28571 |

Table 1: Raw results data
| Date | New Cases | New Deaths | Total Cases | Total Deaths | Est Total Cases |
| --- | --- | --- | --- | --- | --- |
| 23/02/2020 | 62 | 2 | 79 | 2 | 20585.71429 |
| 24/02/2020 | 53 | 0 | 132 | 2 | 25871.42857 |
| 25/02/2020 | 97 | 4 | 229 | 6 | - |
| 26/02/2020 | 93 | 5 | 322 | 11 | - |
| 27/02/2020 | 78 | 1 | 400 | 12 | - |
| 28/02/2020 | 250 | 5 | 650 | 17 | - |
| 29/02/2020 | 238 | 4 | 888 | 21 | - |
| 01/03/2020 | 240 | 8 | 1128 | 29 | - |
| 02/03/2020 | 561 | 6 | 1689 | 35 | - |
| 03/03/2020 | 146 | 17 | 1835 | 52 | - |
| 04/03/2020 | 667 | 28 | 2502 | 80 | - |
| 05/03/2020 | 587 | 27 | 3089 | 107 | - |
| 06/03/2020 | 769 | 41 | 3858 | 148 | - |
| 07/03/2020 | 778 | 49 | 4636 | 197 | - |
| 08/03/2020 | 1247 | 36 | 5883 | 233 | - |
| 09/03/2020 | 1492 | 133 | 7375 | 366 | - |
| 10/03/2020 | 1797 | 98 | 9172 | 464 | - |
| 11/03/2020 | 977 | 167 | 10149 | 631 | - |
| 12/03/2020 | 2313 | 196 | 12462 | 827 | - |
| 13/03/2020 | 2651 | 189 | 15113 | 1016 | - |
| 14/03/2020 | 2547 | 252 | 17660 | 1268 | - |
| 15/03/2020 | 90 | 173 | 17750 | 1441 | - |
| 16/03/2020 | 6230 | 370 | 23980 | 1811 | - |

Figure 3 shows that the model is fairly resistant to varying the case fatality rate from 4-10%.

**Figure 3:**
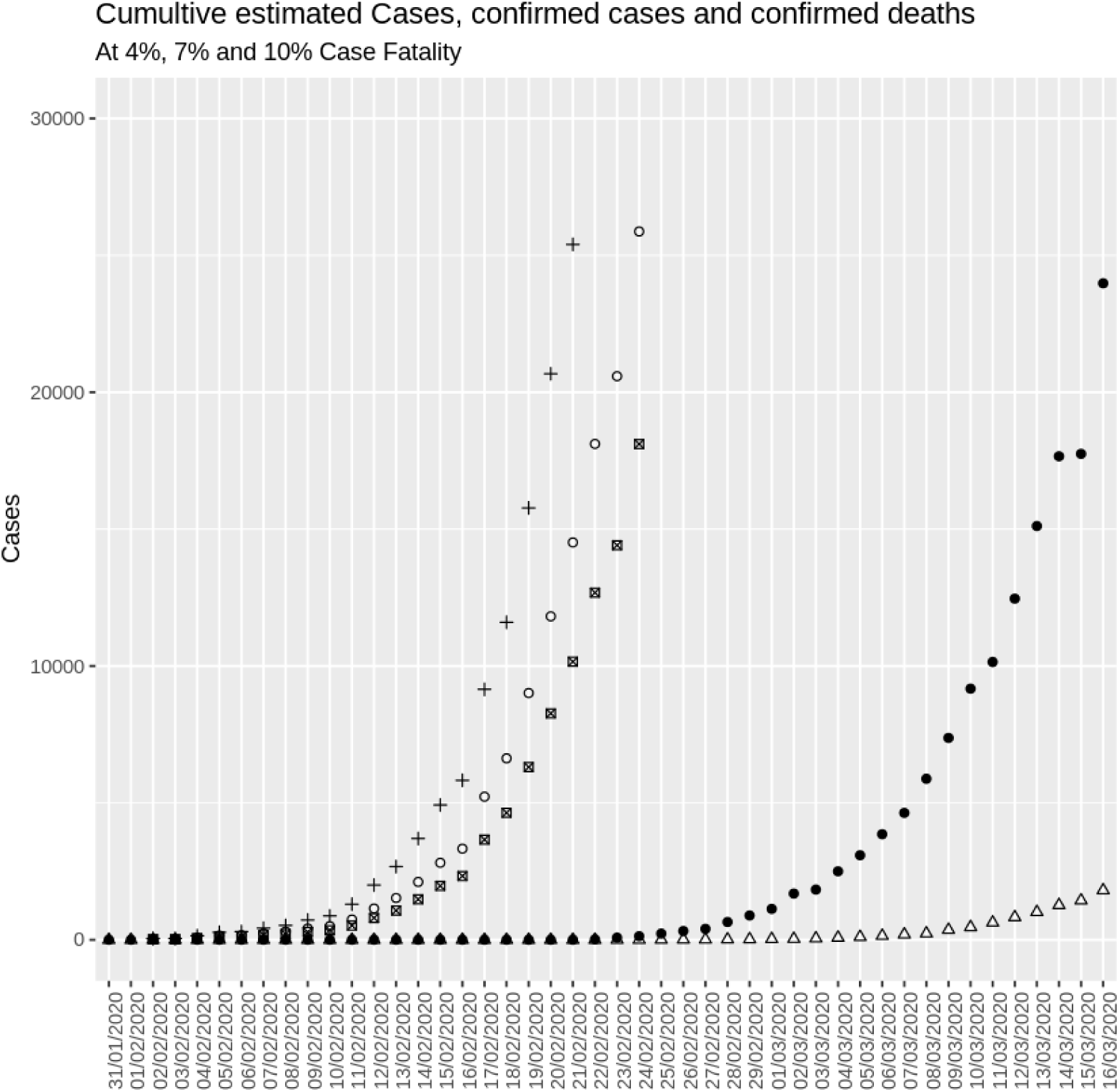
Plotting Estimated Cases at: 4% (crosses), 7% (circles) and 10% (squares), Confirmed Cases (filled triangles), Reported Deaths (triangles).

Italy initially confirmed 3 cases in Italy on the 31^st^ of January 2020, our model predicts in fact there were 28 COVID-19 cases in Italy. From the 31^st^ of January until the 22^nd^ of February there was no detected transmission in Italy and the number confirmed cases remained at 3.

On the 24th of February when our prediction data ends there were 132 cases confirmed by Italian authorities but our model predicts there were near 26000 cases in reality. With less than 0.5% of active cases being confirmed with testing.

Our model predicts that in this period there was undetected transmission resulting in a rise in cases from 28 to 18000. With a doubling period of about 2.5 days.

### Discussion

The large disparity with estimated cases being much higher than confirmed cases indicates that could indicate that increasing majority of cases are not being detected. There seems to have been a period of several weeks where COVID-19 was transmitted in the Italian population undetected. Only a minority of cases appear to be confirmed at any point in time. The model seems robust to a error in case fatality rate.

## Conclusion

In this paper we present evidence that the currently confirmed cases of COVID-19 are a dramatic underestimate of the true point prevalence in Italy and a method to estimate cases from daily deaths of COVID-19. Increasing the mortality would reduce the estimated prevalence but this alone could not make the estimates agree with the confirmed cases in order of magnitude due to our models robustness to varying case fatality ratio in the region of current estimates. This methodology would be applicable to many other conditions and relies only on accurate estimate of deaths due to the condition which can easily be confirmed post-mortem and case mortality (when lower end estimates are above 2%). Without incubation date or data on disease progression an accurate estimate can still be produced but will not provide temporal information but could be used to estimate the time from infection to death.

## Limitations

This model used the spot daily reported deaths which may lag the true date of death due to delays in confirming and then reporting causes of death if COVID-19 was not diagnoses ante mortem. Our estimate of point prevalence varies proportionally to the error in deaths. Deaths due to infection not reported will cause an underestimate in prevalence. Estimating true mortality rates is difficult, and our estimate varies with the reciprocal of the error in the mortality rate. Underestimating mortality will lead to increase in predicted cases. As shown in Figure 3 due to this our model is relatively robust for an error in case fatality ratio 4–10%, which is comparable to reported case fatality ratio estimates. This is an evolving pandemic and due to the long incubation and time to death periods only a short period of point prevalence’s can be estimated. Especially during the incubation period it’s possible for those infected with SARS-CoV2 to travel into Italy, meaning cases predicted may not have acquired the infection in Italy.

## Competing Interests

The authors declare no competing interests.

## Data Availability

Data not included in the article is available on request.

## Acknowledgements

I would like to thank the healthcare workers around the globe for their tireless efforts fighting COVID-19.

## Notes

### Competing Interest Statement

The authors have declared no competing interest.

### Funding Statement

no external funding was received

